# Effects of Dipeptidyl Peptidase-4 Inhibitors and Sulfonylureas on Cognitive and Physical Function in Nursing Home Residents

**DOI:** 10.1101/2021.08.31.21262941

**Authors:** Andrew R. Zullo, Matthew S. Duprey, Robert J. Smith, Roee Gutman, Sarah D. Berry, Medha N. Munshi, David D. Dore

## Abstract

**Aims:** Dipeptidyl peptidase-4 inhibitors (DPP4Is) may mitigate hypoglycemia-mediated declines in cognitive and physical functioning compared to sulfonylureas (SUs), yet comparative studies are unavailable among older adults, especially nursing home (NH) residents. We evaluated the effects of DPP4Is versus SUs on cognitive and physical functioning among NH residents.

**Materials and Methods:** This new-user cohort study included long-stay NH residents aged ≥65 years from the 2007-2010 national US Minimum Data Set (MDS) clinical assessments and linked Medicare claims. We measured cognitive decline from the validated 6-point MDS Cognitive Performance Scale, functional decline from the validated 28-point MDS Activities of Daily Living scale, and hospitalizations or emergency department visits for altered mental status from Medicare claims. We compared 180-day outcomes in residents who initiated a DPP4I versus SU after propensity score matching using Cox regression models.

**Results:** The cohort (N=1,784) had a mean (SD) age of 80 (8) years and was 73% female. Approximately 46% had no or mild cognitive impairment and 35% had no or mild functional impairment before treatment initiation. Compared to SU users, DPP4I users had statistically similar 180-day rates of cognitive decline (HR=0.61, 95%CI 0.31-1.19), altered mental status events (HR=0.71, 95%CI 0.39-1.27), and functional decline (HR=0.89, 95%CI 0.51-1.56).

**Conclusions:** Rates of cognitive and functional decline were not markedly reduced among DPP4I users compared to SU users, but the point estimates and lower 95% confidence bounds do not rule out the possibility that DPP4Is result in reduced rates. Larger studies with greater statistical power should resolve this remaining uncertainty.

## INTRODUCTION

Type 2 diabetes mellitus (T2DM) increases the risk of cognitive^1, 2^ and physical functional impairment^3, 4^ in older adults. Little is known about whether individual classes of glucose-lowering drugs affect the risks of adverse neuropsychological or physical functioning.^1, 5^ Understanding the effects of medications on such outcomes is especially important for frail older adults like nursing home (NH) residents, who are at higher risk of decline and who often have limited ability to regain cognitive or physical function. Furthermore, given their limited life expectancy, NH residents often prioritize preserving cognition, functional independence, and quality of life^6^ over disease-specific or longevity outcomes.^7^

Though the exact mechanisms responsible for T2DM-associated cognitive impairment are unknown, hyperinsulinemia, hyperglycemia, and hypoglycemia may cause cognitive impairment.^8, 9^ Hypoglycemia occurs frequently among NH residents and increases the risk of delirium, impaired cognition, fatigue, weakness, and in severe cases, seizures and unconsciousness.^10-14^ Hyperglycemia also frequently occurs in the NH population, and it can also lead to cognitive and functional impairments similar to those from hypoglycemia.^15, 16^ These symptoms of hypoglycemia and hyperglycemia may result in irreversible decline in cognitive functioning, physical functioning, and quality of life for frail and vulnerable older adults.^17-20^

Dipeptidyl peptidase-4 inhibitors (DPP4Is) and sulfonylureas (SUs) are two of the most commonly prescribed glucose-lowering drug classes for T2DM among NH residents.^21-23^ These oral glucose-lowering agents may improve cognition and the ability to perform activities of daily living by improving blood glucose control, potentially even reversing impaired neuro-motor function.^2, 24-28^ DPP4Is and SUs may differentially affect cognitive and physical functioning through their effects on hypoglycemia and hyperglycemia.^26-28^ SUs are associated with a greater risk of hypoglycemia than DPP4Is.^23, 26-28^ The use of DPP4Is may also reduce amyloid beta accumulation in the brain, which could further help to maintain or slow cognitive decline.^29^ However, hyperglycemia may occur more often with DPP4Is versus SUs. Studies that directly compare cognitive and physical functioning outcomes for DPP4I and SU users are scarce.^24, 30^

We estimated the effects of DPP4Is versus SUs on cognitive functioning, altered mental status events, and physical functioning among frail older NH residents. We hypothesized that DPP4I use would be associated with a lower rate of cognitive impairment, smaller number of altered mental status events, and better physical functioning than SU use because of the comparatively lower rate of hypoglycemia from DPP4Is.^23^

## MATERIALS AND METHODS

### Study Design and Data Source

This was a retrospective new-user cohort study that used the following linked national datasets for the years 2007 – 2010: 100% Medicare fee-for-service enrollment information, Part A inpatient claims, and Part B outpatient claims; 20% Part D prescription drug claims; 100% Minimum Data Set (MDS) version 2.0 assessment records; and 100% Online Survey Certification and Reporting System (OSCAR) data. The MDS is a comprehensive, clinical assessment instrument used to document health status of NH residents, including functional status, psychological, and cognitive status information.^31^ MDS assessments occur at a minimum of every 3 months, and are conducted more frequently for residents with a major recent change in clinical status. OSCAR data provided NH-level information. The study was designed to mimic the hypothetical target trial detailed in Supplementary Table S1.^32^ This study was approved by the Brown University Institutional Review Board.

### Study Population

The study population was adults aged ≥65 years who were long-stay NH residents (>100 days in the NH) on January 1, 2008, or who became a long-stay resident between January 1, 2008 and September 30, 2010. The index date (time zero) was the date of the first eligible dispensing of a DPP4I or SU between January 1, 2008 and September 30, 2010 after becoming a long-stay resident. Data from July 1, 2007 to December 31, 2007 was available to ascertain prior glucose-lowering treatment use. All individuals were required to have one year of continuous enrollment in Medicare fee-for-service Parts A, B, and D immediately preceding the index date. Cohort exclusions are shown in Supplementary Figure S1 and Table S1.

### Exposures and Causal Contrast of Interest

Exposures of interest were new use of DPP4Is (saxagliptin, sitagliptin) or SUs (glimepiride, glipizide, glyburide) in NHs. New use was defined as the first Part D claim for a DPP4I or SU after 6 months without a dispensing for any glucose-lowering treatment other than metformin. As metformin is generally first-line therapy but is commonly contraindicated in the NH setting^33^, its use was permitted and adjusted for but not required for inclusion. The causal contrast of interest was defined as the effect of initiating DPP4Is versus SUs regardless of subsequent treatment discontinuation or switching (i.e., the observational study analog of the intention-to-treat [ITT] estimand).

### Outcomes

The outcomes were decline in physical functioning, decline in cognitive functioning, and hospitalizations or emergency department (ED) visits for altered mental status. We defined physical functional decline as an increase of 3 points on a validated 28-point scale of independence in activities of daily living (ADLs) between the pre-initiation baseline and any available MDS assessment following initiation, up to 180 days after initiation.^34^ A 3-point increase corresponds to a major loss of independence in 1 ADL or incremental loss in 2 or more ADLs.^35^ Cognitive decline was defined as an increase of 1 point on the validated 6-point MDS Cognitive Performance Scale (CPS). A 1-point cognitive decline is considered clinically significant.^36^ Lastly, we defined a hospitalization or ED visit for altered mental status using International Classification of Diseases, Ninth Revision, Clinical Modification (ICD-9-CM) codes 780.0, 780.02, 780.09, or 780.97 in any position on a Part A inpatient hospital claim or a Part B ED claim, though inclusion of ED visits contributed only a small number of additional cases. While no validated algorithm exists for altered mental status events in claims, our definition is consistent with prior literature.^37^ We analyzed each outcome individually. We also explored a pre-specified composite outcome: time to functional or cognitive decline (3-point ADL or 1-point CPS).

### Follow-Up

The start of follow-up (baseline or time zero) for each individual was the day of the first DPP4I or SU dispensing and continued until the earliest event of the following: Medicare disenrollment (from Parts A, B, or D), enrollment in a health maintenance organization (Medicare Advantage), death, an outcome (each evaluated separately), or administrative end of follow-up (September 30, 2010 or 180 days of follow-up). We chose a 180-day outcome period because it is long enough to be clinically meaningful, but short enough that many of these highly vulnerable residents have not yet died. Death is a prevalent competing event that complicates interpretation of longer-term outcomes in the NH setting.

### Baseline characteristics

We identified 190 baseline characteristics that could potentially confound the relationship between receiving DPP4Is versus SUs and the outcomes (Supplementary Table S2). All variables were pre-specified and measured using the most recent available record on or before the index date (time zero).^21, 22, 33^ These variables were obtained from Medicare claims and MDS v2.0 data, which has well-established reliability and validity for assessing the clinical status of NH residents. The MDS v2.0 also provided data on other patient characteristics including pre-treatment physical function^34^ and cognitive status, geriatric syndromes, nutrition, social characteristics, care preferences, and a mortality risk index: Changes in Health, End-stage Disease, and Symptoms Scale (CHESS) score.^38^ We used the OSCAR data to evaluate a variety of NH facility characteristics such as staffing, resident mix, and quality indicators (Supplementary Table S2).

### Statistical Analyses

We adjusted for potential confounding by baseline covariates by estimating the propensity scores using a logistic regression that included 190 baseline characteristics to predict the initiation of DPP4Is versus SUs. We matched DPP4I to SU users on the propensity scores using a 1:1 greedy (nearest neighbor) 5-to-1 digit matching algorithm without replacement.^39^ Cox regression models with robust standard errors to account for clustering within the matched sets were used to estimate hazard ratios (HRs) and 95% confidence intervals (CIs) comparing DPP4I versus SU users.^40^ No covariates were included in the model. Testing for violations of the proportional hazards assumption was unnecessary because we interpret the HRs as a weighted average of the true HRs over the entire follow-up period.^41^ We used SAS, version 9.4 (SAS Institute Inc) for data processing, and Stata version 14.0 (Stata Corp., College Station, TX) and R version 3.4.4 (The R Foundation) for data analyses.

### Stability Analyses

We conducted several stability analyses to test the robustness of our treatment effect estimates to study design and analysis decisions. First, we evaluated more substantial declines by re-defining functional decline as a 4-point decrease and cognitive decline as a 2-point decrease along with a composite outcome of time to larger functional declines (4-point ADL or 2-point CPS). Second, to assess the impact of missing data, we performed multiple imputation with chained equations to impute missing covariate data for 582 residents excluded from the primary analysis. Third, we estimated the propensity scores using generalized boosted regression models to evaluate whether our parametric propensity score estimation model might have been mis-specified. Fourth, we used Fine and Gray competing risks regression models to account for the potential competing risk of death. At the end of each follow-up period, subjects were classified as alive without one of the outcomes of interest, having had the outcome documented in the MDS or claims in that period, or having died without evidence of an outcome event. Lastly, because metformin is often the preferred first-line medication used to treat T2DM, we restricted our study cohort to individuals using metformin at baseline and performed the analyses in this subpopulation.

## RESULTS

### Cohort

Before matching, the initial cohort included 903 new DPP4I users and 6,075 new SU users, and these groups had the same average baseline values of ADL (mean=16) and CPS scores (mean=3). DPP4I initiators had a higher medication burden, used other glucose-lowering treatments more frequently, and had more angiotensin receptor blocker, clopidogrel, fibrate, and omega-3 fatty acid medication use in the prior 12 months than new SU users (Table 1 and Supplementary Table S2). Additionally, DPP4I users were more likely to have abnormal laboratory results, ischemic heart disease, and a prior ED visit for hyperglycemia, but less likely to have a do not resuscitate order.

**Table 1.** Characteristics of Nursing Home Residents Initiating Dipeptidyl Peptidase-4 Inhibitors or Sulfonylureas Before and After Propensity Score Matching.

| Characteristic | n (%) |  |  |  |
| --- | --- | --- | --- | --- |
|  | Before matching |  | After matching |  |
|  | DPP4I users<br>(N=903) | SU users<br>(N=6,075) | DPP4I users<br>(N=892) | SU users<br>(N=892) |
| Age, mean (SD) years | 81 (8) | 81 (8) | 81 (8) | 80 (8) |
| Female sex | 652 (72) | 4,282 (71) | 644 (72) | 661 (74) |
| Race |  |  |  |  |
| White | 651 (72) | 4,593 (76) | 648 (73) | 628 (70) |
| Black | 145 (16) | 963 (16) | 142 (16) | 155 (17) |
| Other | 107 (12) | 519 (8) | 102 (11) | 109 (13) |
| Conditions |  |  |  |  |
| Renal disease | 97 (11) | 572 (9) | 96 (11) | 96 (11) |
| Peripheral vascular disease | 157 (17) | 1,015 (17) | 156 (18) | 164 (18) |
| Heart failure | 284 (32) | 1,744 (29) | 281 (32) | 272 (31) |
| Ischemic heart disease | 162 (18) | 927 (15) | 159 (18) | 153 (17) |
| Depression | 509 (56) | 3,399 (56) | 504 (57) | 483 (54) |
| Chronic Obstructive Pulmonary Disease | 192 (21) | 1,223 (20) | 191 (21) | 202 (23) |
| Hypertension | 692 (77) | 4,681 (77) | 683 (77) | 710 (80) |
| Hip fracture | 15 (2) | 108 (2) | 15 (2) | 25 (3) |
| Number of conditions, median (Q1, Q3) | 6 (4, 7) | 5 (4, 7) | 6 (4, 7) | 6 (4, 8) |
| ADL score, mean (SD)<sup>†</sup> | 16 (8) | 16 (8) | 16 (8) | 17 (7) |
| CPS score, mean (SD)<sup>‡</sup> | 3 (2) | 3 (2) | 3 (2) | 3 (2) |
| CHESS score, mean (SD)<sup>§</sup> | 1 (1) | 1 (1) | 1 (1) | 1 (1) |
| Number of medications, mean (SD) | 14 (5) | 13 (5) | 14 (5) | 14 (5) |
| Medication use |  |  |  |  |
| Metformin | 330 (37) | 1,768 (29) | 325 (36) | 301 (34) |
| Statins | 401 (44) | 2,362 (39) | 394 (44) | 390 (44) |
| Clopidogrel | 168 (19) | 849 (14) | 163 (18) | 164 (18) |
| Warfarin | 141 (16) | 899 (15) | 139 (16) | 137 (15) |
| Antipsychotics | 283 (31) | 1,779 (29) | 281 (32) | 255 (29) |
| Steroids (oral) | 110 (12) | 751 (12) | 109 (12) | 122 (14) |
| Angiotensin-converting enzyme inhibitors | 371 (41) | 2,297 (38) | 368 (41) | 379 (43) |
| Angiotensin receptor blockers | 140 (16) | 682 (11) | 135 (15) | 136 (15) |
| Beta blockers | 455 (50) | 2,698 (44) | 449 (50) | 457 (51) |
| Length of nursing home stay before treatment initiation, median (Q1, Q3) days | 623 (275, 1,260) | 586 (270, 1,177) | 628 (276, 1,263) | 604 (274, 1,248) |
| Any physician visits in prior two weeks | 531 (59) | 3,518 (58) | 519 (58) | 536 (60) |
| Number of physician order changes in prior two weeks, mean (SD) | 2 (2) | 2 (2) | 2 (2) | 2 (2) |
| Any overnight hospitalizations in prior 90 days | 287 (32) | 1,792 (29) | 282 (32) | 295 (33) |
| Any ED visits in prior 90 days | 71 (8) | 490 (8) | 70 (8) | 75 (8) |
| **Nursing Home Facility Characteristics** |  |  |  |  |
| Ownership |  |  |  |  |
| For-profit | 657 (73) | 4,317 (71) | 648 (73) | 626 (70) |
| Non-profit | 193 (21) | 1,341 (22) | 192 (22) | 210 (24) |
| Government | 53 (6) | 417 (7) | 52 (6) | 56 (6) |
| Quality indicators |  |  |  |  |
| % of residents restrained, median (Q1, Q3) | 2 (0, 5) | 2 (0, 5) | 2 (0, 5) | 2 (0, 5) |
| No. of quality-of-life deficiencies, mean (SD) | 4 (6) | 4 (6) | 4 (6) | 4 (7) |
| % of residents with pressure sores, mean (SD) | 7 (4) | 7 (4) | 7 (4) | 7 (5) |
| Staffing |  |  |  |  |
| Direct care hours/resident/day, mean (SD) | 3 (1) | 3 (1) | 3 (1) | 3 (1) |

Abbreviations: DPP4I, dipeptidyl Peptidase-4 Inhibitors; SU, sulfonylureas; SD, standard deviation; Q1, first quartile; Q3, third quartile; ADL, Activities of Daily Living; CPS, Cognitive Performance Scale; CHESS, Changes in Health, End-stage disease and Symptoms and Signs Score; ED, emergency department.
†Physical function was measured using activities of daily living (ADL) via the Minimum Data Set Morris 28-point ADL score. The ADL scores range from 0 to 28, with 0 indicating total independence and 28 indicating total dependence in all ADLs.
‡Cognitive status was measured using the Minimum Data Set Cognitive Performance Scale (CPS), where higher values indicate greater cognitive impairment. The CPS scores range from 0 to 6, with 0 indicating intact cognitive function and 6 indicating very severe cognitive impairment.
§ The Minimum Data Set Changes in Health, End-stage disease and Symptoms and Signs (CHESS) score is a composite
measure addressing changes in health, end-stage disease, and symptoms and signs of medical problems. The CHES scores range from 0 to 5, with 0 indicating no health instability and 5 indicating very high health instability.

Propensity score matching yielded a cohort of 892 new DPP4I users and an equal number of SU users (N=1,784; Table 1). The mean age (SD) was 80 (SD=8) years. Approximately 73% of the cohort was female and 72% was White race, and 46% had no or mild cognitive impairment (CPS score 0-2) and 35% had no or mild functional impairment (ADL score 0-14) before DPP4I or SU initiation. The matching procedure balanced covariates closely^42^ with all but 2 variables having absolute standardized mean differences of 0.06 or less (Supplementary Table S2). The propensity score distribution between new initiators of DPP4Is and of SUs overlapped substantially before and after matching (Supplementary Figure S2).

### Effects on Outcomes

The rates of cognitive decline and altered mental status were not statistically different between DPP4I users and SU users (Table 2, Figure 1, Supplementary Table S3). The rate of cognitive decline among DPP4I users was 0.61 (95%CI 0.31-1.19) times that of SU users, and the rate of an altered mental status event was 0.71 (95%CI 0.39-1.27) times that of SU users (Table 2, Figure 2). DPP4I users were as likely as SU users to have a physical functional decline at 180 days after treatment initiation (HR=0.89, 95%CI 0.51-1.56). The rates of the composite outcome of cognitive or functional decline were not statistically different between DPP4I user and SU users (180 days, HR=0.83, 95%CI 0.53-1.31).

**Table 2.**
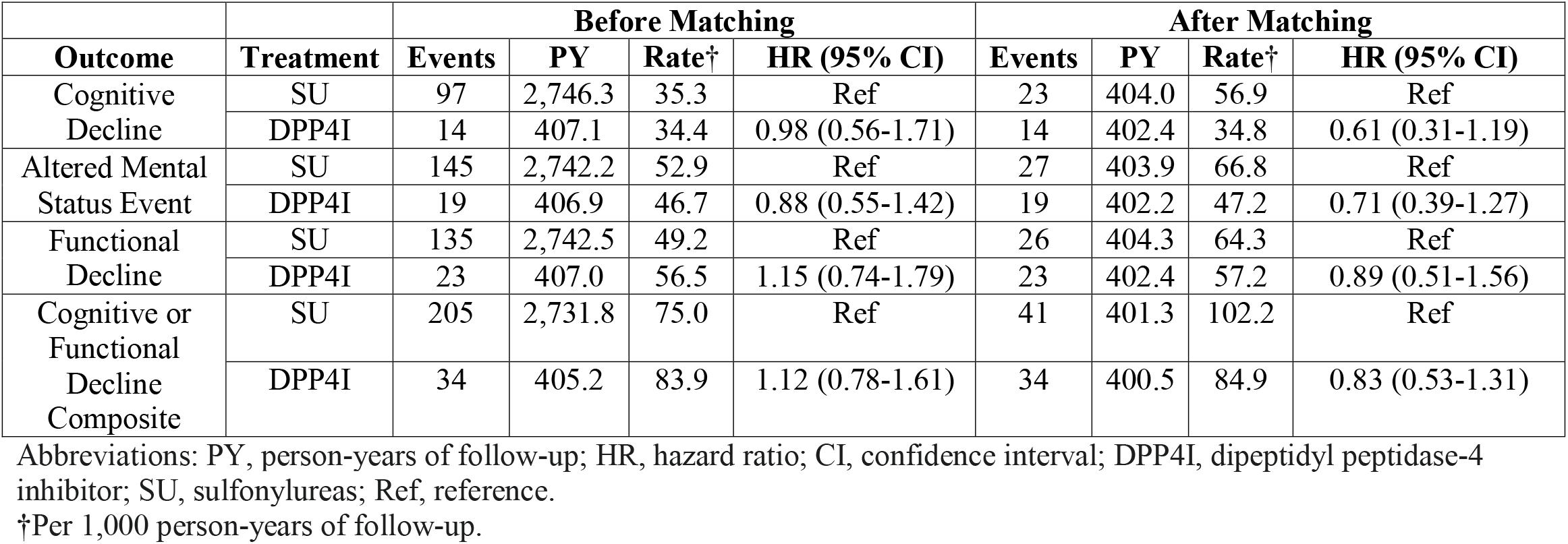
Effects of Dipeptidyl Peptidase-4 Inhibitors (n=892) versus Sulfonylureas (n=892) on 180-Day Outcomes Before and After Propensity Score Matching among Nursing Home Residents.

|  |  | Before Matching |  |  |  | After Matching |  |  |  |
| --- | --- | --- | --- | --- | --- | --- | --- | --- | --- |
| Outcome | Treatment | Events | PY | Rate<sup>†</sup> | HR (95% CI) | Events | PY | Rate<sup>†</sup> | HR (95% CI) |
| Cognitive Decline | SU | 97 | 2,746.3 | 35.3 | Ref | 23 | 404.0 | 56.9 | Ref |
|  | DPP4I | 14 | 407.1 | 34.4 | 0.98 (0.56-1.71) | 14 | 402.4 | 34.8 | 0.61 (0.31-1.19) |
| Altered Mental Status Event | SU | 145 | 2,742.2 | 52.9 | Ref | 27 | 403.9 | 66.8 | Ref |
|  | DPP4I | 19 | 406.9 | 46.7 | 0.88 (0.55-1.42) | 19 | 402.2 | 47.2 | 0.71 (0.39-1.27) |
| Functional Decline | SU | 135 | 2,742.5 | 49.2 | Ref | 26 | 404.3 | 64.3 | Ref |
|  | DPP4I | 23 | 407.0 | 56.5 | 1.15 (0.74-1.79) | 23 | 402.4 | 57.2 | 0.89 (0.51-1.56) |
| Cognitive or Functional Decline Composite | SU | 205 | 2,731.8 | 75.0 | Ref | 41 | 401.3 | 102.2 | Ref |
|  | DPP4I | 34 | 405.2 | 83.9 | 1.12 (0.78-1.61) | 34 | 400.5 | 84.9 | 0.83 (0.53-1.31) |

Abbreviations: PY, person-years of follow-up; HR, hazard ratio; CI, confidence interval; DPP4I, dipeptidyl peptidase-4 inhibitor; SU, sulfonylureas; Ref, reference.
<sup>†</sup>Per 1,000 person-years of follow-up.

**Figure 1.**
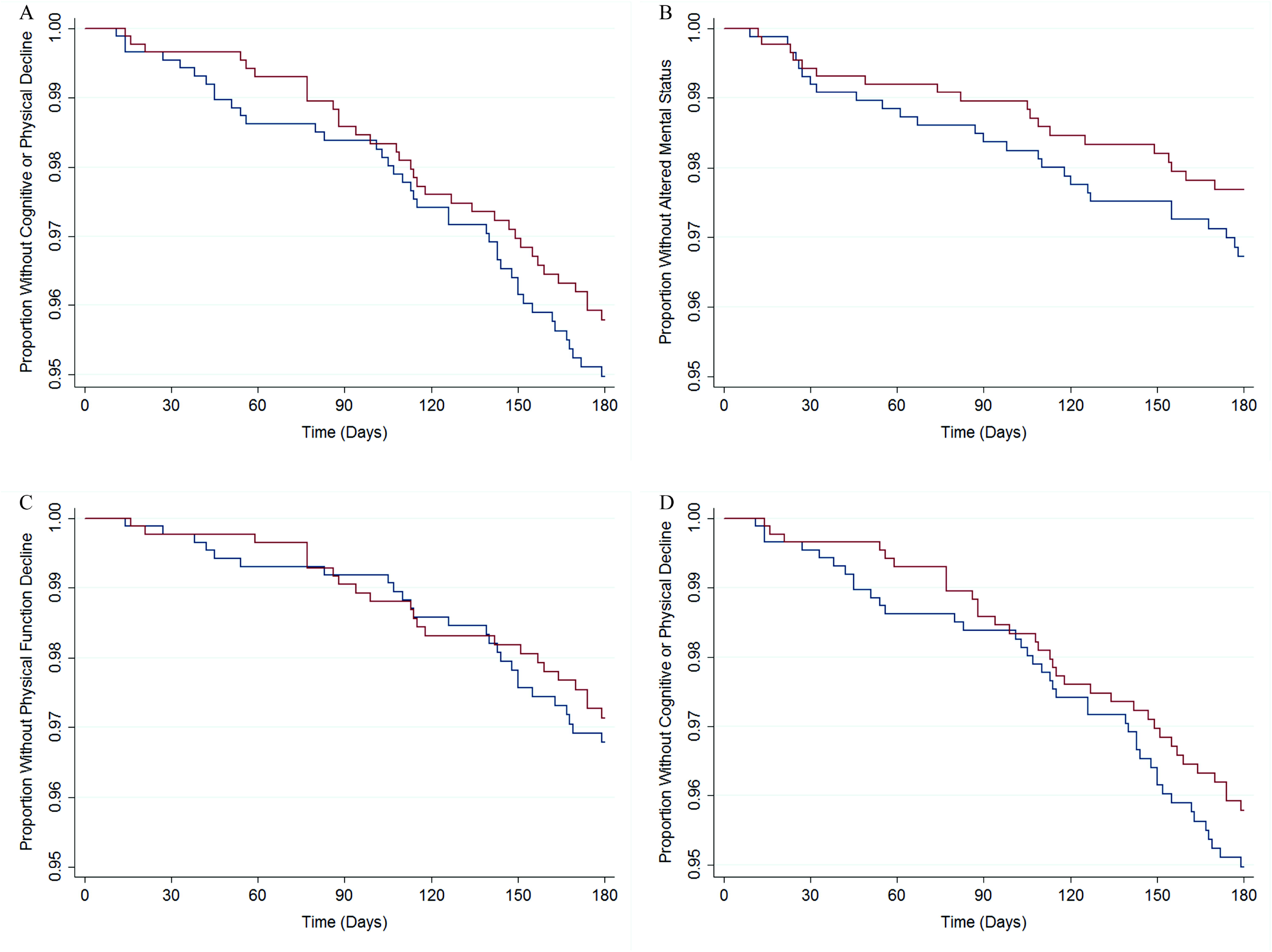
Survival Curves for Cognitive Decline, Altered Mental Status Event, Functional Decline, and Cognitive or Functional Decline Composite Outcomes over 180 Days of Follow-up Stratified by Dipeptidyl Peptidase-4 Inhibitor versus Sulfonylurea Use after Propensity Score Matching among Nursing Home Residents. Panel A shows time to cognitive decline among DPP4I users and SU users. Panel B shows time to hospitalization or emergency department visit for altered mental status. Panel C shows time to functional decline. Panel D shows time to the first of either a cognitive or functional decline. Red lines are DPP4I users; blue lines are SU users. Please refer to Supplementary Table S3 for the corresponding risk table for each survival curve.

**Figure 2.**
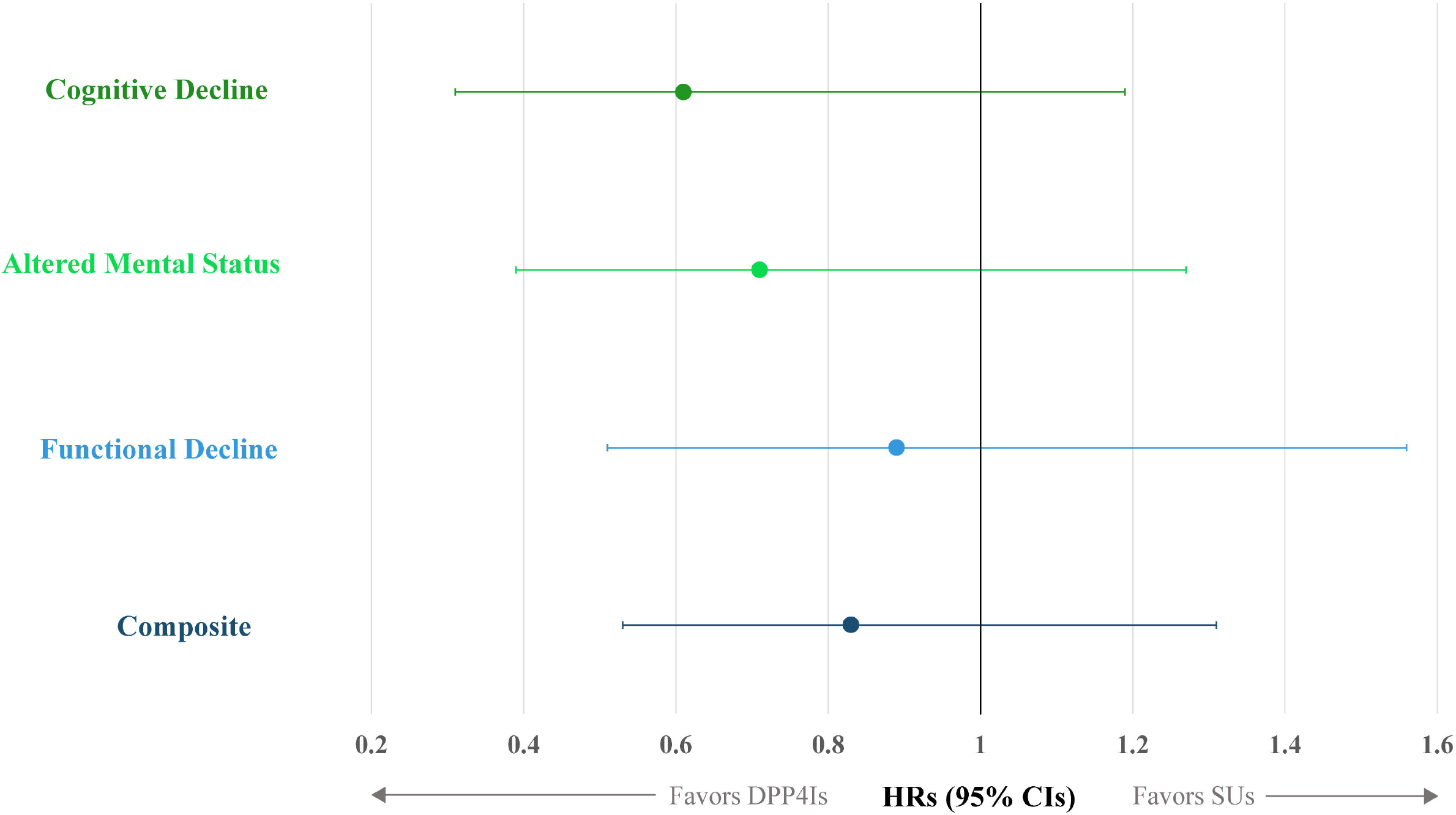
Effects of Dipeptidyl Peptidase-4 Inhibitor versus Sulfonylurea Use on 180-day Outcomes After Propensity Score Matching among Nursing Home Residents. Note: Composite denotes a composite outcome of time to either cognitive or functional decline. Abbreviations: HR, hazard ratio; CI, confidence interval; DPP4I, dipeptidyl peptidase-4 inhibitor; SU, sulfonylurea.

### Stability Analyses

Analyses re-defining cognitive and functional decline using thresholds representing larger changes (2-point increase in CPS score or 4-point increase in the Morris ADL scale) resulted in similar point estimates with decreased precision for the individual outcomes as well as the composite that reflected the most severe measures of cognitive or functional decline (Supplementary Table S4). The main results were generally similar when implementing multiple imputation of missing baseline covariate information (Supplementary Table S5), estimating the propensity score using generalized boosted regression models (Supplementary Table S6), employing Fine and Gray regression models to account for the competing risk of death (Supplementary Table S7), and restricting to metformin users (Supplementary Table S8).

## DISCUSSION

In this national cohort study of NH residents, we observed statistically insignificant differences in rates of cognitive or functional decline and altered mental status among new users of DPP4Is compared to new users of sulfonylureas. However, the point estimates and lower 95% confidence bounds do not exclude the possibility that DPP4I use decreases the rate of functional decline, especially cognitive decline. Providers should not preferentially prescribe DPP4Is to affect change in cognition based on the insufficient evidence currently available, and should instead focus on other side effects for which there is more evidence, such as hypoglycemia.^23^

Few studies have examined the comparative effects of T2DM medications on cognition, physical functioning, or quality of life.^14^ Of the studies that have been conducted, major differences in the study designs and methods preclude any meaningful summative conclusions, especially because just one of those studies evaluated DPP4Is versus SUs.^14, 24^ Rizzo et al. evaluated the effect of DPP4Is versus SUs on changes in cognitive function in older Italian patients with mild cognitive impairment.^24^ The study was not conducted in NH residents and did not appear to explicitly adjust for confounding. The investigators concluded that DPP4I use improved cognitive function compared to SU use.

The closest comparable evidence on physical functioning is a large Veterans Affairs study of the relationship between glycemic control and functional decline (defined as a 2-point increase in the MDS ADL score) in NH residents.^1^ The investigators did not compare treatments, but conducted an analysis stratified by prevalent glucose lowering treatment use—insulin, sulfonylureas, or other glucose-lowering medications. They found that 238 (11.4%) of 2,094 users of glucose-lowering treatments other than sulfonylureas or insulin and 404 (14.1%) of 2,873 sulfonylurea users had a functional decline. However, their analysis did not include adjustment for differences in all relevant factors between these glucose lowering treatments.

Our study has several limitations. Because it is an observational study without randomization of treatments, we cannot rule out the possibility of residual confounding. Our study may have benefited from laboratory information on hemoglobin A1c and other measures of glycemic control that may have influenced prescribers’ decision-making, but direct measures of this information were unavailable. Nonetheless, we obtained good balance on a wide array of baseline covariates between treatment groups, including variables that may be correlates of glycemic control. Moreover, DPP4Is and SUs have a similar place in therapy for treatment of T2DM, which reduces the likelihood of confounding by indication or T2DM severity.^43^

Another important consideration is the size of our final study population and precision of our estimates. Functional status changes were somewhat rare over the follow-up period, and DPP4I use was not common during the study period, resulting in imprecise estimates of effects. These circumstances mandate a more nuanced interpretation of the range of plausible effect estimates, rather than simply seeking to reject or fail to reject the null hypothesis using p-values (i.e., rather than assessing whether estimates are or are not statistically significant). In addition to contributing to the imprecision of our estimates through a reduction in sample size, excluding individuals with a recent history of glucose-lowering treatment use other than metformin may have reduced the generalizability of our findings. Future work in a study population with a larger number of new DPP4I users appears to be merited.

Lastly, there may be under-ascertainment of outcomes. It is possible that declines in cognitive or physical functioning are not noticed or documented by the NH staff between MDS assessments. However, provided that the under-ascertainment is non-differential by treatment group and the specificity of the MDS-based measures is high, relative effect measures are unlikely to be biased. Since the MDS requirements do not differ by glucose-lowering treatment status, differential ascertainment of declines in cognitive and physical functioning is unlikely.

## Conclusion

In summary, the rates of cognitive decline, altered mental status events, and functional decline were not statistically different between DPP4I users and SU users, but the point estimates and lower 95% confidence bounds do no not rule out the possibility that DPP4Is result in reduced rates of cognitive decline among NH residents. Given that the evidence is currently insufficient, providers should not preferentially prescribe DPP4Is to influence cognition.^44^ Given the absence of clinical trials that include NH residents, additional studies using data from routine practice are needed to inform patient-centered treatment decisions for NH residents with T2DM. Because many NH residents, their families, their caregivers, and their clinicians value maintaining functioning and quality of life more than extending lifespan or minimizing traditional clinical adverse events, real world evidence could help guide the selection of T2DM treatments to achieve those goals.

## Data Availability

The data employed in our study are subject to a data use agreement with the Centers for Medicare and Medicaid Services and cannot be made available to other researchers.

## Author Contributions

1. Conception and design: ARZ, RJS, RG, DDD.
2. Acquisition of data: ARZ, DDD.
3. Analysis of data: ARZ, RG.
4. Interpretation of data: All authors.
5. Drafting the article: ARZ, MSD.
6. Revising it critically for important intellectual content: All authors.
7. Final approval of the version to be published: All authors.

