## Supplementary for "Effects of Dipeptidyl Peptidase-4 Inhibitors and Sulfonylureas on Cognitive and Physical Function in Nursing Home Residents"

**Supplementary Table S1.** Summary of the Protocol of the Hypothetical Target Trial Emulated to Compare the Effects of Dipeptidyl Peptidase-4 Inhibitors and Sulfonylureas on Cognitive and Functional Outcomes among Older Nursing Home Residents.

**Supplementary Table S2.** Covariates included in Propensity Score Estimation Model and Covariate Balance Assessed Using Standardized Differences in Means and Proportions Before and After Propensity Score Matching.

**Supplementary Table S3.** Survival Curve Risk Tables for Cognitive Decline, Altered Mental Status, Functional Decline, and Cognitive or Functional Decline Composite Outcomes over 180 Days of Follow-up Stratified by Dipeptidyl Peptidase-4 Inhibitor versus Sulfonylurea Use after Propensity Score Matching among Nursing Home Residents.

**Supplementary Table S4.** Results from Stability Analyses Using a 2-point (Rather Than 1-point) Threshold for the Minimum Data Set Cognitive Performance Scale Score to Identify Cognitive Decline and a 4-point (Rather Than 3-poin) Threshold for the Minimum Data Set Activities of Daily Living Long Form Scale Score to Identify Functional Decline Before and After Propensity Score Matching.

**Supplementary Table S5.** Results from Stability Analyses Using Multiple Imputation of Missing Baseline Covariate Information Before and After Propensity Score Matching.

**Supplementary Table S6.** Results from Stability Analyses Using Generalized Boosted Regression Models to Estimate the Propensity Score.

**Supplementary Table S7.** Results from Stability Analyses using Fine and Gray Regressions to Account for the Competing Risk of Death.

**Supplementary Table S8.** Results from Stability Analyses Restricting to Only Users of Metformin in the Six Months Prior to Time Zero Before† and After Propensity Score Matching.

**Supplementary Figure S1.** Flow Diagram of Study Cohort Creation.

**Supplementary Figure S2.** Propensity Score Distributions by Treatment Group Before and After Matching.

**Supplementary Table S1.** Summary of the Protocol of the Hypothetical Target Trial Emulated to Compare the Effects of Dipeptidyl Peptidase-4 Inhibitors and Sulfonylureas on Cognitive and Functional Outcomes among Older Nursing Home Residents.

| **Protocol Component** | **Description** |
| --- | --- |
| Eligibility criteria | - Nursing home residents aged ≥65 years who reside in the nursing home for more than 100 days. - Nursing home residents must have been continuously enrolled in Medicare Parts A, B, and D and not enrolled in a health maintenance organization (Medicare Advantage) for the 12 months before follow-up begins. - Nursing home residents are ineligible if they are comatose, paralyzed, have cancer, or are in hospice in the 12 months before follow-up begins. - Nursing home residents are ineligible if they use any dose of any glucose-lowering treatment other than metformin in the 6 months before follow-up begins. |
| Treatment strategies | - Initiate any dose of one dipeptidyl peptidase-4 inhibitor drug after a washout of 6 months or longer for both dipeptidyl peptidase-4 inhibitors and sulfonylureas. - Initiate any dose of one sulfonylurea drug after a washout of 6 months or longer for both dipeptidyl peptidase-4 inhibitors and sulfonylureas. - *Note:* Under both treatment strategies, the decision to discontinue or initiate any additional therapies is left to the patient and their healthcare professionals’ discretion. |
| Assignment procedures | - Unblinded random assignment of participants to either treatment strategy at baseline; participants are aware of the treatment strategy that they are assigned to. |
| Follow-up period | - Time zero of follow-up is the time at which a participant meets all of the eligibility criteria (when a person is randomized [assigned] to one of the treatment strategies—dipeptidyl peptidase-4 inhibitor use or sulfonylurea use— at baseline). - The follow-up period of interest is 180 days after time zero. - Follow-up ends at earliest occurrence of an outcome event, (disenrollment from Medicare Parts A, B, or D or enrollment in health maintenance organization [Medicare Advantage] insurance), death, or administrative end of follow-up (September 30, 2010 or 180 days after time zero). |
| Outcomes | - Decline in physical function recorded by a nurse as an increase of 3 points from baseline on the 28-point Minimum Data Set Activities of Daily Living (MDS-ADL) Long Form Scale - Decline in cognitive function recorded by a nurse as an increase of 1 point on the 6-point Minimum Data Set Cognitive Performance Scale (MDS CPS) - Hospitalization or emergency department visit for altered mental status - Composite of decline in physical function of 3 points on the MDS-ADL scale or decline in cognitive function of 1 point on the MDS CPS sale |
| Causal contrasts | - Intention-to-treat eﬀect   - The effect of being assigned to any dose of dipeptidyl peptidase-4 inhibitor at baseline versus being assigned to any dose of sulfonylurea at baseline, regardless of whether participants adhere to their assigned treatment strategy during follow-up (regardless of treatment discontinuation or switching) |
| Analysis plan | - Participants will be analyzed “as randomized” to the treatment strategies. - Estimated from an inverse probability weighted Cox regression model adjusted for covariates measured at time zero (baseline). - Inverse probability weights will be estimated as a function of 190 characteristics measured at time zero, including measures of cognitive status, functional status, geriatric syndromes, metformin use, demographics, medical conditions, healthcare utilization, nutrition, care preferences, social function, and the nursing home facility. |

**Supplementary Table S2.** Covariates included in the Propensity Score Estimation Model and Covariate Balance Assessed Using Standardized Differences in Means and Proportions Before and After Propensity Score Matching.

|  |  | Original Value | | Absolute Value | |
| --- | --- | --- | --- | --- | --- |
| # | Label | Before Matching | After Matching | Before Matching | After Matching |
| 1 | Number of comorbidities | 0.06 | -0.01 | 0.06 | 0.01 |
| 2 | Morris activities of daily living scale (0-28 point) | 0.02 | -0.03 | 0.02 | 0.03 |
| 3 | Facility: total nurse aide full time equivalents per 100 beds | -0.02 | -0.01 | 0.02 | 0.01 |
| 4 | Facility: total physical therapy full time equivalents per 100 beds | -0.02 | 0.04 | 0.02 | 0.04 |
| 5 | Facility: total registered nurse full time equivalents per 100 beds | 0.05 | -0.01 | 0.05 | 0.01 |
| 6 | Facility: pharmacist full time equivalents | -0.02 | 0.03 | 0.02 | 0.03 |
| 7 | Facility: Percentage of residents physically restrained | -0.02 | 0.05 | 0.02 | 0.05 |
| 8 | Facility: medication error rate | -0.03 | -0.03 | 0.03 | 0.03 |
| 9 | Facility: count of quality-of-life deficiencies | 0.03 | -0.02 | 0.03 | 0.02 |
| 10 | Facility: Percentage of residents physically restrained | 0.02 | 0.01 | 0.02 | 0.01 |
| 11 | Facility: Percentage of residents covered by Medicaid insurance | 0.05 | 0.03 | 0.05 | 0.03 |
| 12 | Facility: Percentage of residents on a pharmacy pain management program | -0.03 | -0.02 | 0.03 | 0.02 |
| 13 | Facility: Percentage of residents receiving antianxiety medications | 0.02 | 0.03 | 0.02 | 0.03 |
| 14 | Facility: Percentage of residents receiving antidepressants | -0.06 | 0.05 | 0.06 | 0.05 |
| 15 | Facility: Percentage of residents with bedsores | 0.05 | -0.02 | 0.05 | 0.02 |
| 16 | Facility: acuity of residents (acuity index) | 0.05 | 0.00 | 0.05 | 0.00 |
| 17 | Facility: Percentage of residents with advance directives | -0.05 | -0.01 | 0.05 | 0.01 |
| 18 | Medication Burden (Number of medications) | 0.24 | 0.03 | 0.24 | 0.03 |
| 19 | Duration of nursing home stay before sulfonylurea or dipeptidyl peptidase-4 inhibitor initiation | 0.02 | -0.01 | 0.02 | 0.01 |
| 20 | Number of orders changed by physician | 0.11 | -0.03 | 0.11 | 0.03 |
| 21 | Number of physician visits | 0.05 | -0.05 | 0.05 | 0.05 |
| 22 | Female sex | -0.04 | 0.04 | 0.04 | 0.04 |
| 23 | Race/ethnicity | 0.13 | 0.05 | 0.13 | 0.05 |
| 24 | Age at index treatment initiation | 0.11 | 0.05 | 0.11 | 0.05 |
| 25 | Marital status | 0.06 | 0.04 | 0.06 | 0.04 |
| 26 | Highest educational attainment | 0.08 | 0.05 | 0.08 | 0.05 |
| 27 | Primary language spoken, English or non-English | 0.07 | 0.05 | 0.07 | 0.05 |
| 28 | Customary routine includes usual attendance at church, temple, synagogue, or other place of worship | -0.07 | -0.04 | 0.07 | 0.04 |
| 29 | Body mass index at index treatment (DPP4I or SU) initiation | 0.15 | 0.03 | 0.15 | 0.03 |
| 30 | Diabetes mellitus | 0.10 | 0.00 | 0.10 | 0.00 |
| 31 | Hyperthyroidism | 0.04 | 0.05 | 0.04 | 0.05 |
| 32 | Hypothyroidism | 0.00 | 0.05 | 0.00 | 0.05 |
| 33 | Ischemic heart disease | 0.07 | 0.02 | 0.07 | 0.02 |
| 34 | Arrhythmias | -0.03 | -0.02 | 0.03 | 0.02 |
| 35 | Heart failure | 0.06 | 0.02 | 0.06 | 0.02 |
| 36 | Deep vein thrombosis | 0.00 | -0.02 | 0.00 | 0.02 |
| 37 | Hypotension | -0.01 | -0.02 | 0.01 | 0.02 |
| 38 | Hypertension | -0.01 | -0.06 | 0.01 | 0.06 |
| 39 | Peripheral vascular disease | 0.02 | -0.02 | 0.02 | 0.02 |
| 40 | Other (non-ischemic) cardiovascular disease | -0.01 | 0.03 | 0.01 | 0.03 |
| 41 | Arthritis | 0.02 | 0.00 | 0.02 | 0.00 |
| 42 | Missing limb | -0.03 | -0.04 | 0.03 | 0.04 |
| 43 | Osteoporosis | 0.01 | -0.04 | 0.01 | 0.04 |
| 44 | Bone fracture (other than hip) | -0.02 | -0.03 | 0.02 | 0.03 |
| 45 | Alzheimer’s disease | -0.02 | -0.01 | 0.02 | 0.01 |
| 46 | Aphasia | 0.00 | 0.05 | 0.00 | 0.05 |
| 47 | Stroke | 0.01 | -0.05 | 0.01 | 0.05 |
| 48 | Non-Alzheimer’s dementia | -0.03 | 0.00 | 0.03 | 0.00 |
| 49 | Antibiotic-resistant infection | 0.01 | 0.00 | 0.01 | 0.00 |
| 50 | Respiratory infection | 0.01 | 0.00 | 0.01 | 0.00 |
| 51 | Seizure disorder | 0.03 | 0.03 | 0.03 | 0.03 |
| 52 | Anxiety disorder | -0.02 | -0.01 | 0.02 | 0.01 |
| 53 | Depression | 0.01 | 0.05 | 0.01 | 0.05 |
| 54 | Bipolar disorder | 0.03 | 0.04 | 0.03 | 0.04 |
| 55 | Emphysema/chronic obstructive pulmonary disease | 0.03 | -0.03 | 0.03 | 0.03 |
| 56 | Cataracts | 0.02 | 0.01 | 0.02 | 0.01 |
| 57 | Diabetic retinopathy | -0.01 | 0.06 | 0.01 | 0.06 |
| 58 | Glaucoma | 0.07 | 0.02 | 0.07 | 0.02 |
| 59 | Macular degeneration | 0.00 | 0.01 | 0.00 | 0.01 |
| 60 | Anemia | 0.04 | 0.00 | 0.04 | 0.00 |
| 61 | Renal failure | 0.04 | 0.00 | 0.04 | 0.00 |
| 62 | Urinary tract infection | 0.07 | 0.00 | 0.07 | 0.00 |
| 63 | Wound infection | 0.06 | 0.03 | 0.06 | 0.03 |
| 64 | Any acute episode or a flare-up of a recurrent or chronic health problem | 0.09 | -0.01 | 0.09 | 0.01 |
| 65 | Accidental fall hospitalization | 0.00 | 0.01 | 0.00 | 0.01 |
| 66 | Accidental fall emergency department visit | 0.05 | 0.00 | 0.05 | 0.00 |
| 67 | Intracranial hemorrhage hospitalization | 0.05 | 0.00 | 0.05 | 0.00 |
| 68 | Intracranial hemorrhage emergency department visit | -0.01 | -0.03 | 0.01 | 0.03 |
| 69 | Stroke hospitalization | 0.03 | -0.04 | 0.03 | 0.04 |
| 70 | Stroke emergency department visit | 0.03 | -0.03 | 0.03 | 0.03 |
| 71 | Acute myocardial infarction hospitalization | 0.00 | 0.02 | 0.00 | 0.02 |
| 72 | Acute myocardial infarction emergency department visit | 0.00 | 0.03 | 0.00 | 0.03 |
| 73 | Hypoglycemia hospitalization | 0.11 | -0.06 | 0.11 | 0.06 |
| 74 | Hypoglycemia emergency department visit | 0.12 | 0.04 | 0.12 | 0.04 |
| 75 | Hyperglycemia hospitalization | 0.12 | 0.02 | 0.12 | 0.02 |
| 76 | Hyperglycemia emergency department visit | 0.11 | 0.03 | 0.11 | 0.03 |
| 77 | Hip fracture hospitalization | 0.02 | -0.05 | 0.02 | 0.05 |
| 78 | Hip fracture emergency department visit | 0.00 | -0.03 | 0.00 | 0.03 |
| 79 | Altered mental status hospitalization | 0.01 | 0.03 | 0.01 | 0.03 |
| 80 | Altered mental status emergency department visit | 0.04 | 0.00 | 0.04 | 0.00 |
| 81 | Heart failure hospitalization | 0.12 | -0.01 | 0.12 | 0.01 |
| 82 | Heart failure emergency department visit | 0.09 | 0.01 | 0.09 | 0.01 |
| 83 | Weight gain or loss of 3 or more pounds | -0.05 | 0.02 | 0.05 | 0.02 |
| 84 | Frailty proxy/overall health stability: Changes in Health, End-stage disease, and Signs and Symptoms (CHESS) Scale (0 to 5); 0=Not at all unstable, 5=Highly unstable | 0.07 | 0.05 | 0.07 | 0.05 |
| 85 | Change in ability to perform activities of daily living | 0.03 | 0.05 | 0.03 | 0.05 |
| 86 | Cognitive status (Fries and Morris Cognitive Performance Scale score) | 0.11 | 0.04 | 0.11 | 0.04 |
| 87 | Highest level of pain present in the prior 7 days (i.e., frequency and severity with which resident complains or shows evidence of pain) on last MDS assessment prior to index treatment (DPP4I or SU) initiation | 0.07 | 0.02 | 0.07 | 0.02 |
| 88 | Fell in the past 31 to 180 days on last MDS assessment prior to index treatment (DPP4I or SU) initiation | -0.04 | -0.04 | 0.04 | 0.04 |
| 89 | Change in cognitive status, skills, or abilities as compared to status of 90 days ago or since last assessment if less than 90 days | 0.08 | 0.04 | 0.08 | 0.04 |
| 90 | Cognitive ability varies over time | 0.10 | 0.03 | 0.10 | 0.03 |
| 91 | Bowel incontinence | 0.10 | 0.05 | 0.10 | 0.05 |
| 92 | Bladder incontinence | 0.10 | 0.05 | 0.10 | 0.05 |
| 93 | Pressure ulcers, presence and stage | 0.08 | 0.05 | 0.08 | 0.05 |
| 94 | Dizziness/vertigo in prior 7 days on last MDS assessment prior to index treatment (DPP4I or SU) initiation | -0.01 | -0.03 | 0.01 | 0.03 |
| 95 | Delirium resident assessment triggered | -0.08 | 0.02 | 0.08 | 0.02 |
| 96 | Hallucinations | -0.02 | 0.05 | 0.02 | 0.05 |
| 97 | Hearing aid present and not used regularly | -0.08 | 0.01 | 0.08 | 0.01 |
| 98 | Hearing aid present and used | -0.02 | 0.00 | 0.02 | 0.00 |
| 99 | Side vision problems or decreased peripheral vision | -0.02 | -0.03 | 0.02 | 0.03 |
| 100 | Glasses, contact lenses, or magnifying glass used | -0.06 | -0.02 | 0.06 | 0.02 |
| 101 | Skin tears or cuts (other than surgery) | -0.05 | -0.01 | 0.05 | 0.01 |
| 102 | Edema | 0.01 | -0.01 | 0.01 | 0.01 |
| 103 | Metformin use in the 12 months prior to index treatment (DPP4I or SU) initiation | 0.16 | 0.04 | 0.16 | 0.04 |
| 104 | Intermediate-acting insulin use >6 months but ≤12 months prior to index treatment (DPP4I or SU) initiation | 0.06 | 0.04 | 0.06 | 0.04 |
| 105 | Long-acting insulin use >6 months but ≤12 months prior to index treatment (DPP4I or SU) initiation | 0.24 | 0.00 | 0.24 | 0.00 |
| 106 | Rapid-acting insulin use >6 months but ≤12 months prior to index treatment (DPP4I or SU) initiation | 0.24 | 0.01 | 0.24 | 0.01 |
| 107 | Short-acting insulin use >6 months but ≤12 months prior to index treatment (DPP4I or SU) initiation | 0.16 | 0.02 | 0.16 | 0.02 |
| 108 | Thiazolidinedione use >6 months but ≤12 months prior to index treatment (DPP4I or SU) initiation | 0.21 | -0.01 | 0.21 | 0.01 |
| 109 | Alpha-glucosidase inhibitor use >6 months but ≤12 months prior to index treatment (DPP4I or SU) initiation | 0.06 | 0.02 | 0.06 | 0.02 |
| 110 | Miscellaneous anti-hypertensive use in the 12 months prior to index treatment (DPP4I or SU) initiation | 0.07 | 0.00 | 0.07 | 0.00 |
| 111 | Proton pump inhibitor use in the 12 months prior to index treatment (DPP4I or SU) initiation | 0.14 | -0.02 | 0.14 | 0.02 |
| 112 | Raloxifene use in the 12 months prior to index treatment (DPP4I or SU) initiation | 0.02 | 0.01 | 0.02 | 0.01 |
| 113 | Antiarrhythmic use in the 12 months prior to index treatment (DPP4I or SU) initiation | 0.01 | 0.05 | 0.01 | 0.05 |
| 114 | Angiotensin-converting enzyme inhibitor use in the 12 months prior to index treatment (DPP4I or SU) initiation | 0.07 | -0.03 | 0.07 | 0.03 |
| 115 | Alpha blocker use in the 12 months prior to index treatment (DPP4I or SU) initiation | -0.04 | 0.00 | 0.04 | 0.00 |
| 116 | Benzodiazepine use in the 12 months prior to index treatment (DPP4I or SU) initiation | 0.02 | 0.01 | 0.02 | 0.01 |
| 117 | Angiotensin receptor blocker use in the 12 months prior to index treatment (DPP4I or SU) initiation | 0.13 | 0.00 | 0.13 | 0.00 |
| 118 | Aspirin use in the 12 months prior to index treatment (DPP4I or SU) initiation | -0.03 | 0.00 | 0.03 | 0.00 |
| 119 | Beta blocker use in the 12 months prior to index treatment (DPP4I or SU) initiation | 0.12 | -0.02 | 0.12 | 0.02 |
| 120 | Bile acid resin use in the 12 months prior to index treatment (DPP4I or SU) initiation | 0.04 | 0.02 | 0.04 | 0.02 |
| 121 | Calcium channel blocker use in the 12 months prior to index treatment (DPP4I or SU) initiation | 0.05 | 0.02 | 0.05 | 0.02 |
| 122 | Clopidogrel use in the 12 months prior to index treatment (DPP4I or SU) initiation | 0.13 | 0.00 | 0.13 | 0.00 |
| 123 | Antidepressant use in the 12 months prior to index treatment (DPP4I or SU) initiation | 0.07 | 0.05 | 0.07 | 0.05 |
| 124 | Ezetimibe use in the 12 months prior to index treatment (DPP4I or SU) initiation | 0.04 | 0.03 | 0.04 | 0.03 |
| 125 | Fibrate use in the 12 months prior to index treatment (DPP4I or SU) initiation | 0.12 | 0.06 | 0.12 | 0.06 |
| 126 | Gabapentin or pregabalin use in the 12 months prior to index treatment (DPP4I or SU) initiation | 0.08 | 0.02 | 0.08 | 0.02 |
| 127 | Glucagon use in the 12 months prior to index treatment (DPP4I or SU) initiation | 0.10 | 0.01 | 0.10 | 0.01 |
| 128 | Potassium-sparing diuretic use in the 12 months prior to index treatment (DPP4I or SU) initiation | 0.07 | 0.00 | 0.07 | 0.00 |
| 129 | Long-acting opioid use in the 12 months prior to index treatment (DPP4I or SU) initiation | 0.02 | 0.01 | 0.02 | 0.01 |
| 130 | Mood-stabilizing medication use in the 12 months prior to index treatment (DPP4I or SU) initiation | 0.08 | 0.01 | 0.08 | 0.01 |
| 131 | Muscle relaxant medication use in the 12 months prior to index treatment (DPP4I or SU) initiation | 0.09 | -0.01 | 0.09 | 0.01 |
| 132 | Niacin use in the 12 months prior to index treatment (DPP4I or SU) initiation | 0.00 | 0.03 | 0.00 | 0.03 |
| 133 | Nonbenzodiazepine hypnotic use in the 12 months prior to index treatment (DPP4I or SU) initiation | 0.08 | -0.01 | 0.08 | 0.01 |
| 134 | Omega-3 fatty acid medication use in the 12 months prior to index treatment (DPP4I or SU) initiation | 0.09 | 0.00 | 0.09 | 0.00 |
| 135 | Antipsychotic use in the 12 months prior to index treatment (DPP4I or SU) initiation | 0.04 | 0.06 | 0.04 | 0.06 |
| 136 | Statin use in the 12 months prior to index treatment (DPP4I or SU) initiation | 0.11 | 0.01 | 0.11 | 0.01 |
| 137 | Oral steroid use in the 12 months prior to index treatment (DPP4I or SU) initiation | -0.01 | -0.04 | 0.01 | 0.04 |
| 138 | Thiazide diuretic use in the 12 months prior to index treatment (DPP4I or SU) initiation | 0.06 | 0.00 | 0.06 | 0.00 |
| 139 | Warfarin use in the 12 months prior to index treatment (DPP4I or SU) initiation | 0.02 | 0.01 | 0.02 | 0.01 |
| 140 | Bisphosphonate use in the 12 months prior to index treatment (DPP4I or SU) initiation | -0.02 | -0.03 | 0.02 | 0.03 |
| 141 | Resisted taking medications, activities of daily living assistance, or eating | 0.04 | 0.03 | 0.04 | 0.03 |
| 142 | Number of new medications | 0.05 | 0.01 | 0.05 | 0.01 |
| 143 | Any intravenous medications | 0.07 | 0.04 | 0.07 | 0.04 |
| 144 | Complains about the taste of many foods | 0.08 | 0.04 | 0.08 | 0.04 |
| 145 | Leaves 25% or more of food uneaten at most meals | -0.02 | 0.02 | 0.02 | 0.02 |
| 146 | On a planned weight change program | 0.05 | 0.01 | 0.05 | 0.01 |
| 147 | Parenteral/intravenous feeding | 0.05 | -0.01 | 0.05 | 0.01 |
| 148 | Mechanically altered diet | -0.01 | 0.03 | 0.01 | 0.03 |
| 149 | Therapeutic diet | -0.03 | -0.01 | 0.03 | 0.01 |
| 150 | Dietary supplement between meals | 0.02 | 0.03 | 0.02 | 0.03 |
| 151 | Chewing problems | 0.00 | 0.00 | 0.00 | 0.00 |
| 152 | Nutrition status care plan implemented | 0.09 | 0.00 | 0.09 | 0.00 |
| 153 | Dehydration/fluid status resident assessment triggered | 0.07 | -0.02 | 0.07 | 0.02 |
| 154 | Dehydration/fluid status care plan implemented | 0.07 | -0.01 | 0.07 | 0.01 |
| 155 | Feeding restrictions advanced directive documented in the medical record | -0.05 | 0.02 | 0.05 | 0.02 |
| 156 | Nutritional status resident assessment triggered | 0.07 | 0.02 | 0.07 | 0.02 |
| 157 | Some or all of resident’s natural teeth were lost | 0.04 | 0.01 | 0.04 | 0.01 |
| 158 | Customary routine includes use of tobacco at least daily | -0.03 | 0.04 | 0.03 | 0.04 |
| 159 | Customary routine includes alcoholic beverages at least weekly | -0.02 | 0.05 | 0.02 | 0.05 |
| 160 | Abnormal laboratory values | 0.11 | -0.06 | 0.11 | 0.06 |
| 161 | Hypoglycemia hospitalizations | 0.11 | -0.05 | 0.11 | 0.05 |
| 162 | Hypoglycemia emergency department visit | 0.12 | 0.04 | 0.12 | 0.04 |
| 163 | Transitions between care settings recently | 0.12 | 0.07 | 0.12 | 0.07 |
| 164 | Ostomy (bowel) | -0.06 | 0.00 | 0.06 | 0.00 |
| 165 | Dialysis use | 0.04 | 0.02 | 0.04 | 0.02 |
| 166 | Ventilator or respirator use | 0.02 | 0.02 | 0.02 | 0.02 |
| 167 | Nutrition or hydration intervention to manage skin problems | 0.05 | 0.01 | 0.05 | 0.01 |
| 168 | Received preventative or protective foot care | 0.01 | 0.04 | 0.01 | 0.04 |
| 169 | Evaluation by a licensed mental specialist | 0.04 | 0.03 | 0.04 | 0.03 |
| 170 | Prefers exercise or sports | 0.01 | -0.04 | 0.01 | 0.04 |
| 171 | Do not hospitalize advanced directive documented in the medical record | -0.05 | 0.05 | 0.05 | 0.05 |
| 172 | Do not resuscitate advanced directive documented in the medical record | -0.12 | 0.00 | 0.12 | 0.00 |
| 173 | Significant change in self-sufficiency as compared to status of 90 days ago or since last assessment if less than 90 days | 0.03 | 0.04 | 0.03 | 0.04 |
| 174 | At ease doing self-initiated activities | -0.07 | 0.03 | 0.07 | 0.03 |
| 175 | Established own goals | -0.04 | 0.05 | 0.04 | 0.05 |
| 176 | Communication scale (communication performance) | 0.10 | 0.05 | 0.10 | 0.05 |
| 177 | Change in ability to express, understand, or hear information | 0.03 | 0.02 | 0.03 | 0.02 |
| 178 | Problem behaviors present | -0.02 | 0.04 | 0.02 | 0.04 |
| 179 | Change in behavior status as compared to 90 days ago or since last assessment if less than 90 days | 0.10 | 0.03 | 0.10 | 0.03 |
| 180 | Family participation in resident’s care | 0.05 | 0.05 | 0.05 | 0.05 |
| 181 | Customary routine includes daily contact with relatives or close friends | -0.05 | 0.02 | 0.05 | 0.02 |
| 182 | Family member responsible for resident | 0.00 | -0.03 | 0.00 | 0.03 |
| 183 | Facility: Staff hours per resident | 0.09 | 0.05 | 0.09 | 0.05 |
| 184 | Facility: part of a nursing home chain | -0.02 | 0.02 | 0.02 | 0.02 |
| 185 | Facility: Class of ownership (for-profit, non-profit, government) | 0.05 | 0.05 | 0.05 | 0.05 |
| 186 | Facility: Organized family group | 0.04 | 0.05 | 0.04 | 0.05 |
| 187 | Facility: % of other private pay clients | 0.12 | 0.07 | 0.12 | 0.07 |
| 188 | Calendar year of sulfonylurea or dipeptidyl peptidase-4 inhibitor initiation | 0.15 | 0.04 | 0.15 | 0.04 |
| 189 | Number of emergency department visits | 0.08 | 0.06 | 0.08 | 0.06 |
| 190 | Number of overnight hospitalizations | 0.08 | 0.06 | 0.08 | 0.06 |

**Supplementary Table S3.** Survival Curve Risk Tables for Cognitive Decline, Altered Mental Status, Functional Decline, and Cognitive or Functional Decline Composite Outcomes over 180 Days of Follow-up Stratified by Dipeptidyl Peptidase-4 Inhibitor versus Sulfonylurea Use after Propensity Score Matching among Nursing Home Residents.

|  |  | **Number at Risk During Follow-Up** | | | | | | |
| --- | --- | --- | --- | --- | --- | --- | --- | --- |
| **Outcome** | **Treatment** | **0 days** | **30 days** | **60 days** | **90 days** | **120 days** | **150 days** | **180 days** |
| Cognitive Decline (Panel A) | SU | 892 | 870 | 845 | 822 | 802 | 767 | 724 |
|  | DPP4I | 892 | 873 | 841 | 816 | 795 | 765 | 728 |
| Altered Mental Status (Panel B) | SU | 892 | 868 | 844 | 820 | 801 | 769 | 727 |
|  | DPP4I | 892 | 869 | 841 | 816 | 794 | 767 | 728 |
| Functional Decline (Panel C) | SU | 892 | 871 | 847 | 824 | 802 | 764 | 721 |
|  | DPP4I | 892 | 873 | 843 | 814 | 794 | 767 | 723 |
| Composite of Cognitive or Functional Decline (Panel D) | SU | 892 | 869 | 842 | 818 | 794 | 755 | 710 |
|  | DPP4I | 892 | 872 | 840 | 811 | 789 | 759 | 717 |

Abbreviations: SU, sulfonylureas; DPP4I, dipeptidyl peptidase-4 inhibitor.

**Supplementary Table S4.** Results from Stability Analyses Using a 2-point (Rather Than 1-point) Threshold for the Minimum Data Set Cognitive Performance Scale Score to Identify Cognitive Decline and a 4-point (Rather Than 3-point) Threshold for the Minimum Data Set Activities of Daily Living Long Form Scale Score to Identify Functional Decline Before and After Propensity Score Matching.

|  | **Hazard Ratio (95% Confidence Interval)** | |
| --- | --- | --- |
| **Outcome** | **Unmatched** | **Matched** |
| Cognitive Decline | 0.97 (0.34-2.75) | 0.81 (0.22-3.00) |
| Functional Decline | 1.19 (0.70-2.02) | 0.95 (0.48-1.88) |
| Cognitive or Functional Decline Composite | 1.13 (0.78-1.62) | 0.84 (0.53-1.32) |

**Supplementary Table S5.** Results from Stability Analyses Using Multiple Imputation of Missing Baseline Covariate Information Before† and After Propensity Score Matching.

|  | **Hazard Ratio (95% Confidence Interval)** | |
| --- | --- | --- |
| **Outcome** | **Unmatched** | **Matched** |
| Cognitive Decline | 0.92 (0.53-1.60) | 0.69 (0.33-1.45) |
| Altered Mental Status | 0.81 (0.51-1.31) | 0.64 (0.32-1.29) |
| Physical Functional Decline | 1.21 (0.79-1.85) | 0.94 (0.51-1.74) |
| Cognitive or Functional Decline Composite | 1.13 (0.80-1.61) | 0.89 (0.53-1.49) |
| †Note that multiple imputation was implemented in the full cohort before propensity score matching. | | |

**Supplementary Table S6.** Results from Stability Analyses Using Generalized Boosted Regression Models to Estimate the Propensity Score.

| **Outcome** | **Hazard Ratio†**  **(95% Confidence Interval)** |
| --- | --- |
| Cognitive Decline | 0.71 (0.34-1.45) |
| Altered Mental Status | 0.86 (0.43-1.73) |
| Physical Functional Decline | 0.68 (0.35-1.32) |
| Cognitive or Functional Decline Composite | 0.75 (0.45-1.26) |
| †Results presented are after matching on the propensity score since only the matched results are affected by the alternate estimation of propensity scores using generalized boosted regression models. | |

**Supplementary Table S7.** Results from Stability Analyses using Fine and Gray Regressions to Account for the Competing Risk of Death Before and After Propensity Score Matching.

|  | **Hazard Ratio (95% Confidence Interval)** | |
| --- | --- | --- |
| **Outcome** | **Unmatched** | **Matched** |
| Cognitive Decline | 0.97 (0.53-1.64) | 0.61 (0.30-1.16) |
| Altered Mental Status | 0.88 (0.53-1.38) | 0.88 (0.53-1.38) |
| Physical Functional Decline | 1.15 (0.72-1.75) | 0.88 (0.50-1.55) |
| Cognitive or Functional Decline Composite | 1.12 (0.76-1.58) | 0.83 (0.52-1.30) |

**Supplementary Table S8.** Results from Stability Analyses Restricting to Only Users of Metformin in the Six Months Prior to Time Zero Before† and After Propensity Score Matching.

|  | **Hazard Ratio (95% Confidence Interval)** | |
| --- | --- | --- |
| **Outcome** | **Unmatched** | **Matched** |
| Cognitive Decline | 0.51 (0.16-1.65) | 0.33 (0.07-1.66) |
| Altered Mental Status | 1.19 (0.55-2.57) | 0.87 (0.31-2.41) |
| Physical Functional Decline | 1.36 (0.66-2.83) | 1.00 (0.37-2.69) |
| Cognitive or Functional Decline Composite | 0.81 (0.40-1.64) | 0.61 (0.25-1.49) |
| †Note that the restriction to only metformin users was implemented before estimation of or matching on the propensity score. | | |

**Supplementary Figure S1.** Flow Diagram of Study Cohort Creation.

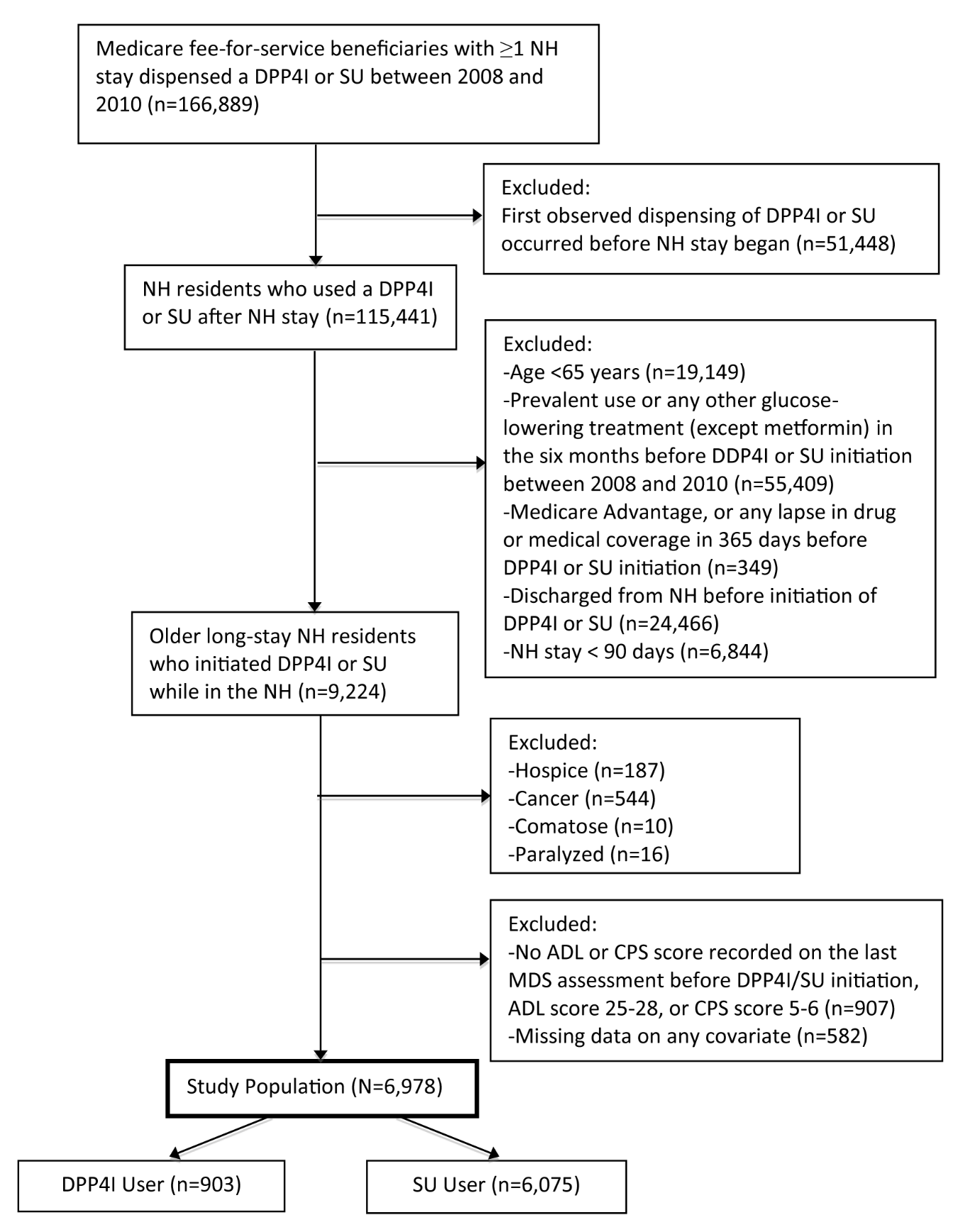

**Supplementary Figure S2.** Propensity Score Distributions by Treatment Group Before and After Propensity Score Matching.

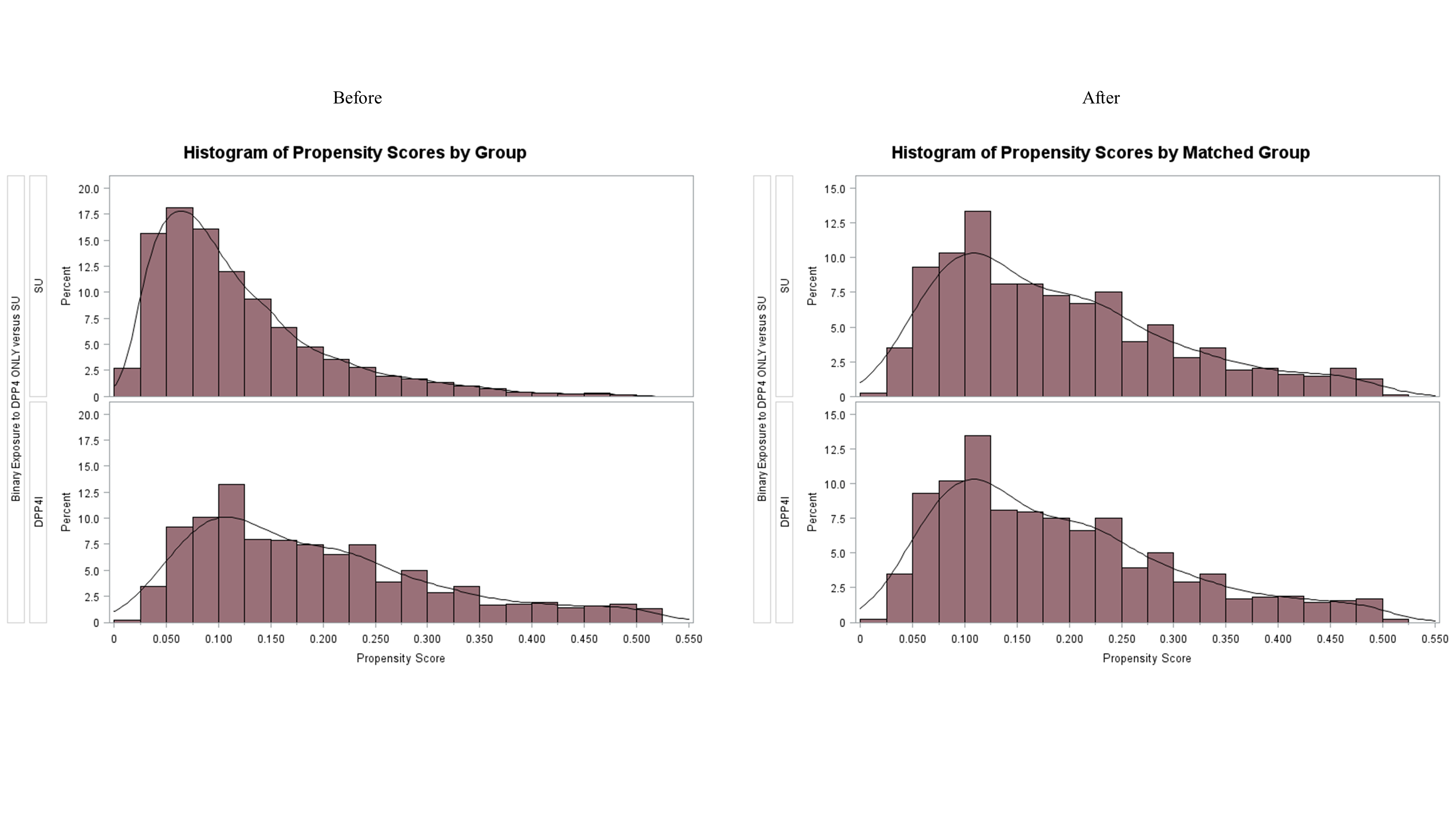
